# When will the battle against novel coronavirus end in Wuhan: a SEIR modeling analysis

**DOI:** 10.1101/2020.02.16.20023804

**Authors:** Kangkang Wan, Jing Chen, Changming Lu, Lanlan Dong, Zhicheng Wu, Lianglu Zhang

## Abstract

Recent outbreak of 2019-nCoV in Wuhan raised serious public health concerns. By February 15, 2020 in Wuhan, the total number of confirmed infection cases has reached 37,914, and the number of deaths has reached 1123, accounting for 56.9% of the total confirmed cases and 73.7% of the total deaths in China. People are eager to know when the epidemic will be completely controlled and when people’s work and life will be on the right track. In this study we analyzed the epidemic dynamics and trend of 2019-nCoV in Wuhan by using the data after the closure of Wuhan city till February 12, 2020 based on the SEIR modeling method. The optimal parameters were estimated as R_0_=1.44 (interquartile range: 1.40-1.47),TI=14 (interquartile range: 14-14) and TE=3.0 (interquartile range: 2.8-3.1). Based on these parameters, the number of infected individuals in Wuhan city may reach the peak around February 19 at about 45,000 people. Once entering March, the epidemic would gradually decline, and end around the late March. It is worth noting that the above prediction is based on the assumption that the number of susceptible population N = 200,000 will not increase. If the epidemic situation is not properly controlled, the peak of infected number can be further increased and the peak time will be a little postponed. It was expected that the epidemic would subside in early March, and disappear gradually towards the late March.

## Introduction

Wuhan is the largest city in central China with a total population of more than 11 million^[1]^. The epidemic of 2019-nCoV pneumonia has been raging in the whole country, especially in Hubei province for nearly a month. In late December 2019, 67 cases of 2019-nCoV pneumonia were reported in Wuhan^[2]^. In order to prevent the further spread of 2019-nCoV, Wuhan began to close the city from 10:00 on January 23, banning all vehicles from entering and leaving the city. Tens of thousands of medical staff, soldiers and people from all walks of life have been involved in the campaign.

The spread of the epidemic has caused a huge threat to people’s health and life safety, at the same time, it has a serious impact on China’s social life and national economy. By February 15, 2020, the total number of confirmed cases has reached 37914, and the number of deaths has reached 1123 in Wuhan, accounting for 56.9% of the total confirmed cases and 73.7% of the total deaths in China^[3]^. With the increase of medical staff from all over the country, the opening of several large novel hospitals, and the adoption of anti epidemic measures, more patients can get efficient and timely treatment. The number of confirmed cases increased sharply on February 12 and 13, while the total number of suspected cases decreased gradually^[3]^.

People are eager to know when the epidemic will be completely controlled and when people’s work and life will be on the right track. In order to help the public to understand the future trend of the epidemic, we analyzed the epidemic dynamic and trend of 2019-nCoV in Wuhan city by using the SEIR modeling method based on the actual data and published references.

## Methods

### 1. Epidemic transmission model

The SEIR model is a classical epidemic model for the flows of people between four states: susceptible (S), exposed (E), infected (I), and recovery (R). Each of those variables represents the number of people in those groups. The relationship among the four groups is elucidated in Figure 1, where *β1* is the probability of S to E after I contacts S, *γ1* is the probability of E to I, and *γ2* is the probability of I to R. Since 2019-nCoV is also infectious in the incubation period, we introduced parameter *β2* here to represent the probability of S to E after E contact S. We used the “susceptible – exposed – infected – recovered” model^[4]^ to describe the prevalent characteristics of 2019-nCoV in Wuhan.

**Fig. 1.**
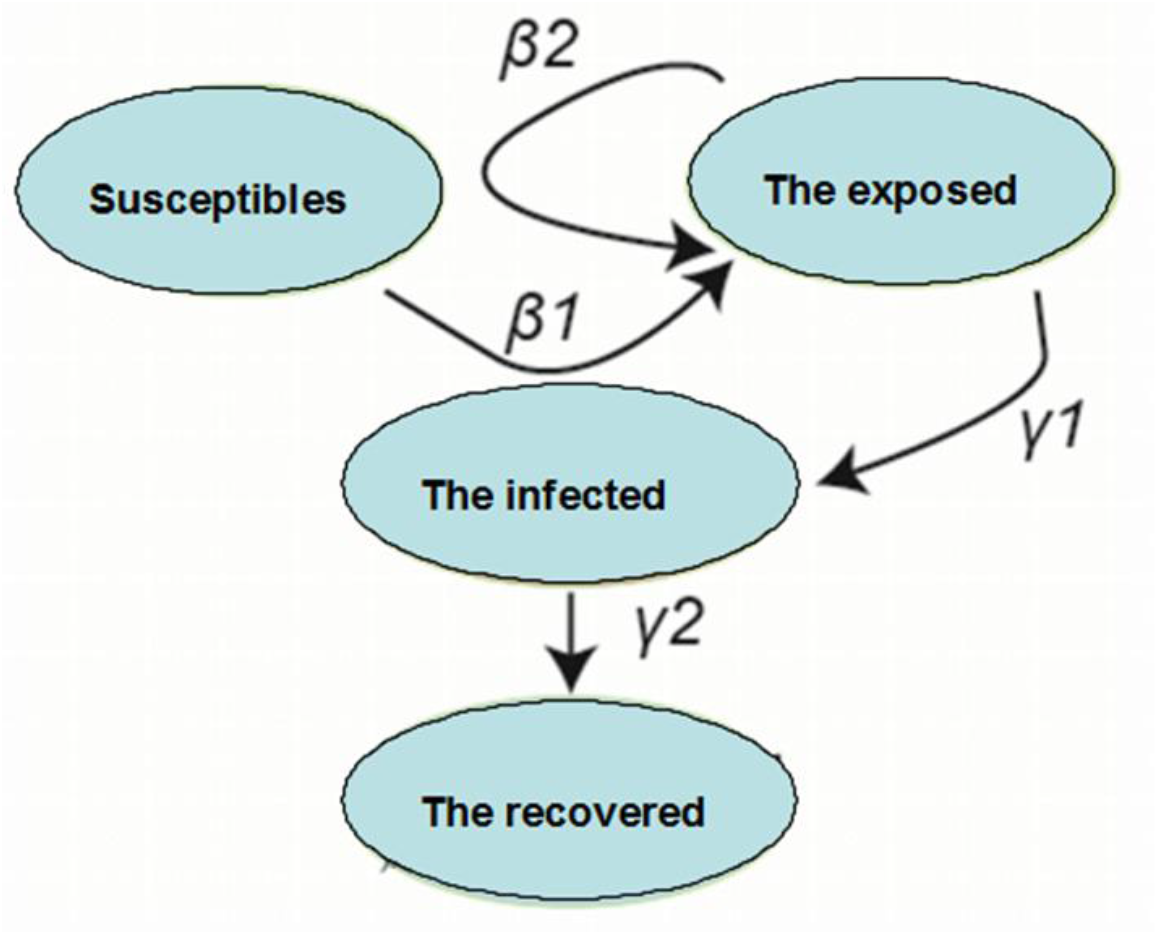
Relationship among four groups according to SEIR model.

This is an ordinary differential equation model, described by the following equations:

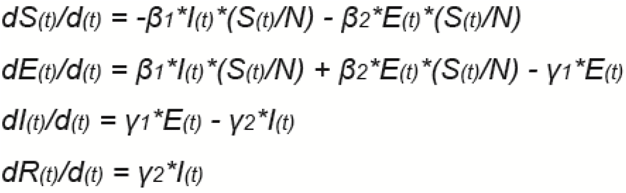

Among which, *S(t), E(t), I(t) and R(t)* represent the number of people in the group of the susceptible, the exposed, the infected and the recovered on the day t, respectively. N is the total number of possible contact people, which is assumed to be fixed and N = S +E + I + R.

The simulation used the fourth-order Runge-Kutta algorithm^[5]^ to solve it numerically, with a step size fixed at 0.01 for R_0_, and at 0.1 for TE and TI.

### 2 Estimation of parameters for the model

Parameters *β1, β2, γ1* and *γ2* were estimated according to the reference^[6]^ using the formula below:

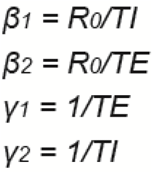

Among which, R_0_ is the basic reproduction number, TI is the time of infectious period, TE is the time of incubation period, and *β1, β2, γ1* and *γ2* share the same meanings as in Figure 1.We set the range of TE to 1-7 days, TI to 1-14 days, R_0_ to 1-5 days according to the reference^[7]^.

The optimized parameters of TE, TI and R_0_ were determined on the condition of RMSE being the smallest.

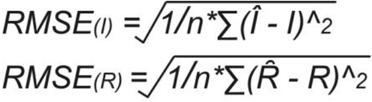

Where I and Î indicate the real and simulated number of the infected, respectively while the R and 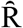 indicate the real and simulated number of the recovery in a specific time, respectively.

Calculate the real number of people (I and R) and the smallest RMSE of Î and 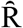 obtained by the model, and choose the TE, TI and R_0_ with the smallest RMSE as the optimal model parameters.

In order to avoid over-fitting in parameter optimization, we took 80% data each time, and repeated this process 100 times, and finally took the average value of R_0_, TI and TE of 100 times as the optimal parameter values.

### 3 Data source

The data were collected from the official website of Hubei Provincial Health Committee (http://wjw.hubei.gov.cn/)^[3]^, and shown in Table 1. We used the data of 22 days from January 22 to February 12 when Wuhan city was shut down and all the public transportation was suspended.

**Table 1.** Number of the infected and the recovery at specific date in Wuhan

| Date | The infected | The recovery |
| --- | --- | --- |
| 22-Jan | 425 | 28 |
| 23-Jan | 495 | 31 |
| 24-Jan | 572 | 39 |
| 25-Jan | 618 | 40 |
| 26-Jan | 698 | 42 |
| 27-Jan | 1,590 | 45 |
| 28-Jan | 1,905 | 75 |
| 29-Jan | 2261 | 82 |
| 30-Jan | 2639 | 103 |
| 31-Jan | 3215 | 139 |
| 1-Feb | 4109 | 171 |
| 2-Feb | 5142 | 224 |
| 3-Feb | 6384 | 303 |
| 4-Feb | 8351 | 368 |
| 5-Feb | 10117 | 431 |
| 6-Feb | 11618 | 534 |
| 7-Feb | 13603 | 698 |
| 8-Feb | 14982 | 877 |
| 9-Feb | 16902 | 1044 |
| 10-Feb | 18454 | 1206 |
| 11-Feb | 19588 | 1377 |
| 12-Feb | 32944 | 1915 |

Source: http://wjw.hubei.gov.cn/

For construction of the model, data of 22 days were divided into two stages. The first stage is from January 23 to February 7, and the second stage is from February 8 to February 12. During the second stage, Wuhan city took a number of measures, including timely diagnosis, timely treatment and effective isolation of the infected population, which will have an important impact on the parameters of the model.

### 4 Initial parameter settings

To establish the model, we firstly estimated the parameters of the susceptible (S), the exposed (E), the infected (I) and the recovery(R) based on the latest data issued on February 12:

N = 200000, which is the total number of potential close contacts in Wuhan on February 12.

S=N-I, in which S is the number of the susceptible and I is the number of the infected.

I_0_ = 425, which is the number of susceptible individuals at the beginning of the model run.

E_0_ = 426, which is the number of exposed individuals at the beginning of the model run

R_0_ = 28, which is the number of recovered individuals at the beginning of the model run

## Results and Analysis

### Epidemic prediction based on SEIR model

The epidemic of the novel coronavirus pneumonia in Wuhan was studied by SEIR modeling. The results showed that, at the time when Wuhan was closed, the number of initially infected individuals was I_0_ = 425, the number of initially exposed individuals was E_0_ = 426, and the number of initially recovered patients was R_0_ = 28.

Next, we separated the data into two stages: January 22-February 7 and February 8-February 12. In the first stage, TI = 14(interquartile range:14-14), TE = 3.0 (interquartile range : 2.8-3.1), R_0_ = 1.44 (interquartile range : 1.40-1.47) (Figure 2). The data showed that the infectious time of the infected person (I) is 14 days, and the incubation period is about 3 days, which is close to the data (Interquartile range: 3.0-7.2) estimated in the reference ^[8]^. The propagation base R_0_ of this study is 1.44, which is significantly lower than the R_0_ estimated by other papers before the closure of Wuhan^[9-11]^.

**Fig 2.**
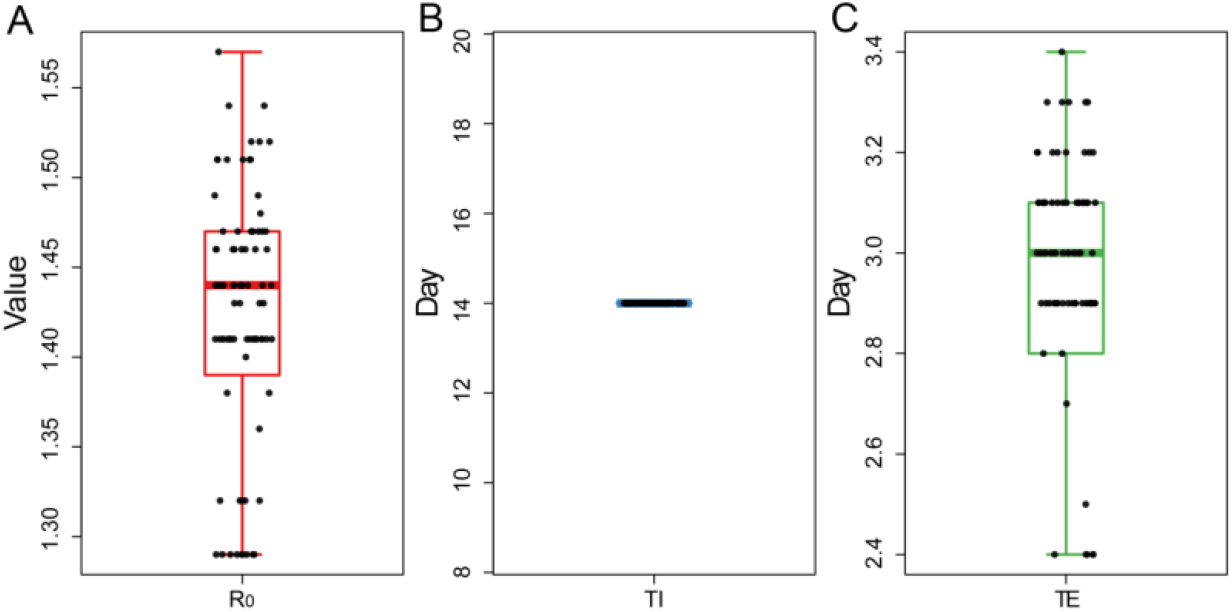
Distribution of the R_0_ (A), TI (B) and TE (C) for 100 random samplings.

In the second stage (after February 8), we set the number of susceptible population N to be fixed at 200000, and the infection cycle of infected population decreased from 14 days to 4 days, i.e. TI = 4, which was estimated according to the data of 5 days from February 8 to February 12, so as to get the epidemic development trend of 90 days since January 22, including the number of infected people, the number of latent people and the number of recovered people (Figure 3, Table 3). The results showed that the number of infected people increased slowly in the early stage (January 22-january 31), but during February 1-February 12, the number of infected people increased rapidly, which is expected to peak around February 19, reaching about 45000 people. Subsequently, the number of infections will decrease. Once entering March, the epidemic would gradually decline, and the epidemic would end around the end of March. It is worth noting that the above prediction is based on the assumption that the number of susceptible population N = 200000 will not increase.

**Table 2.**
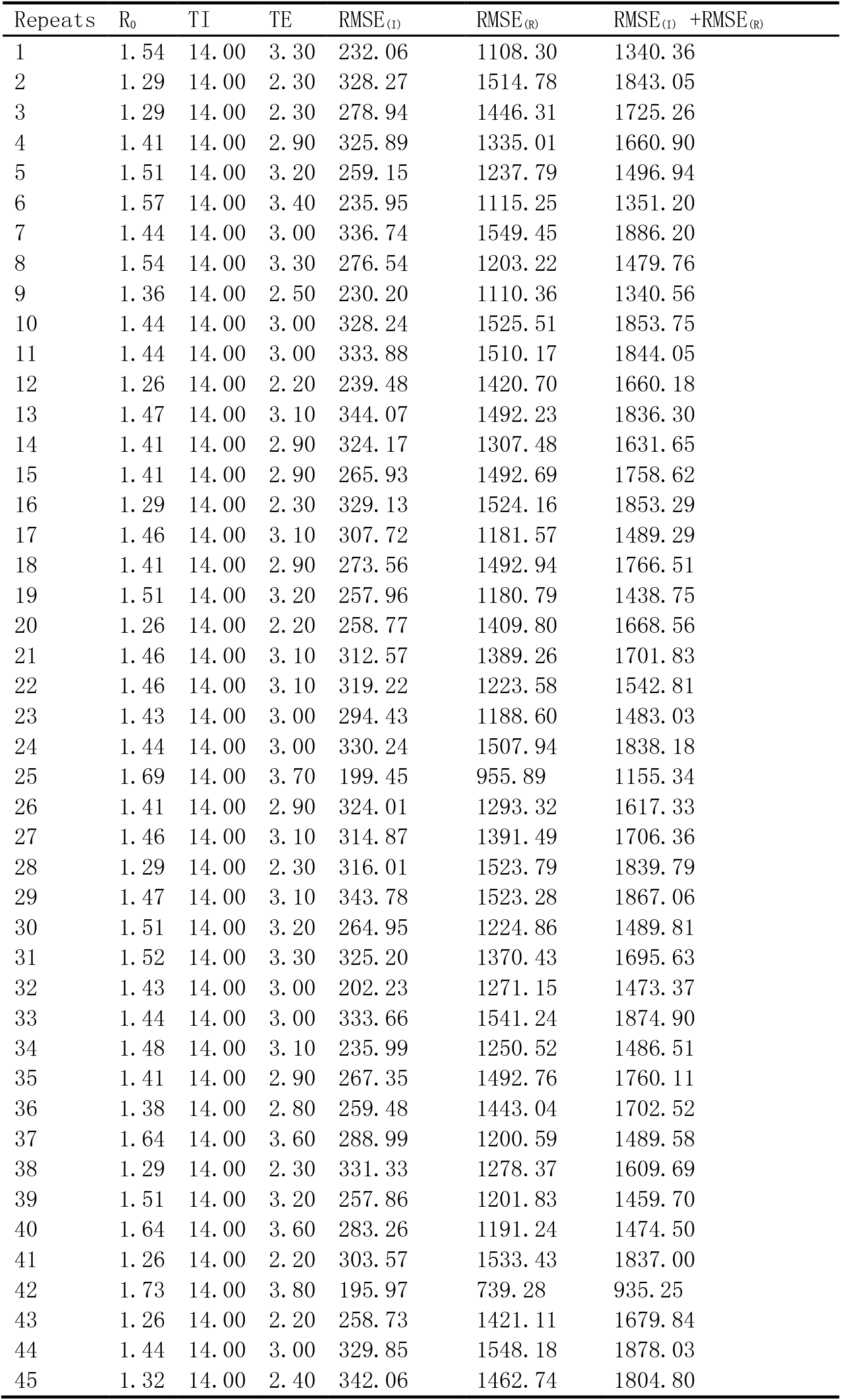
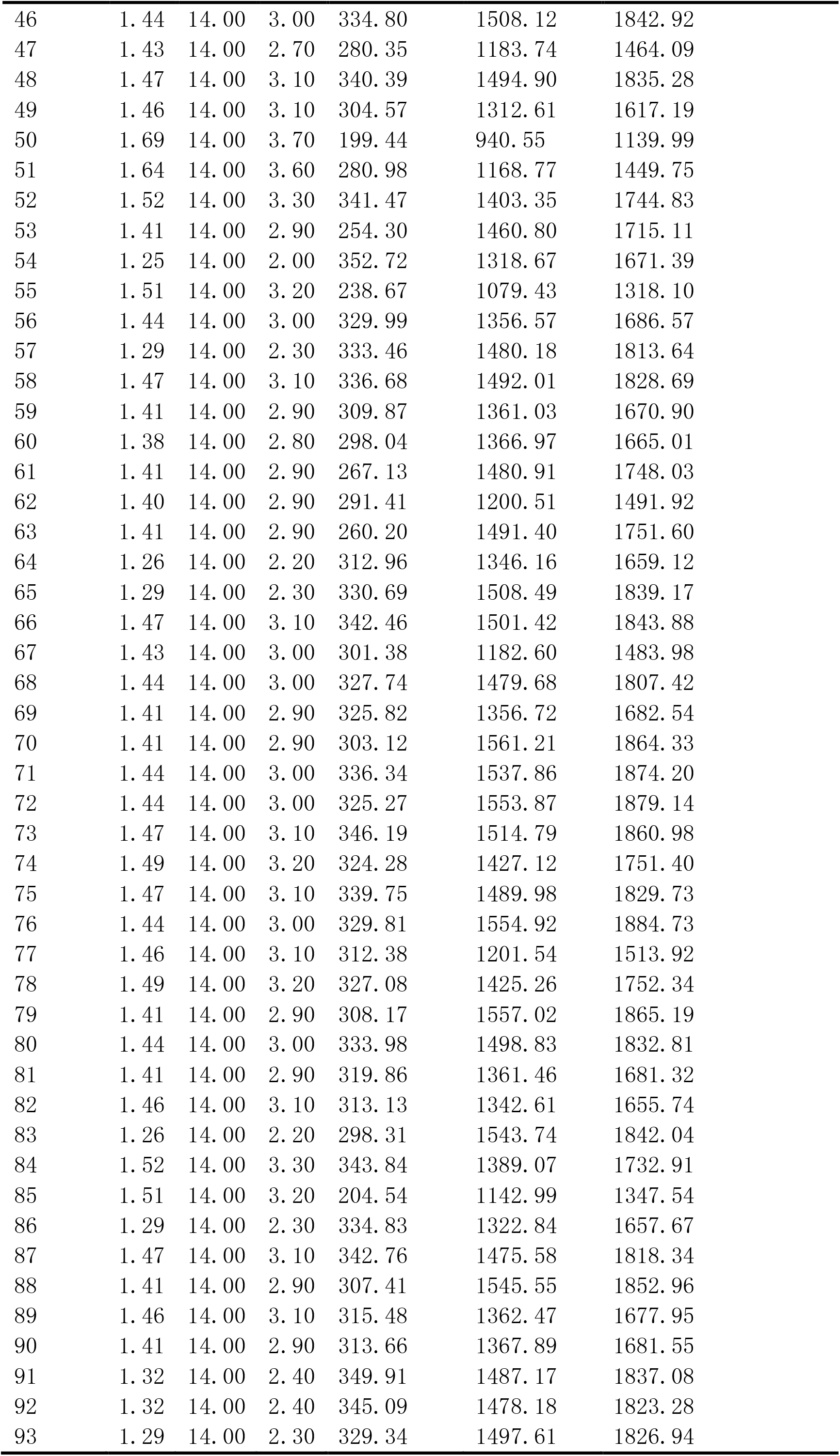
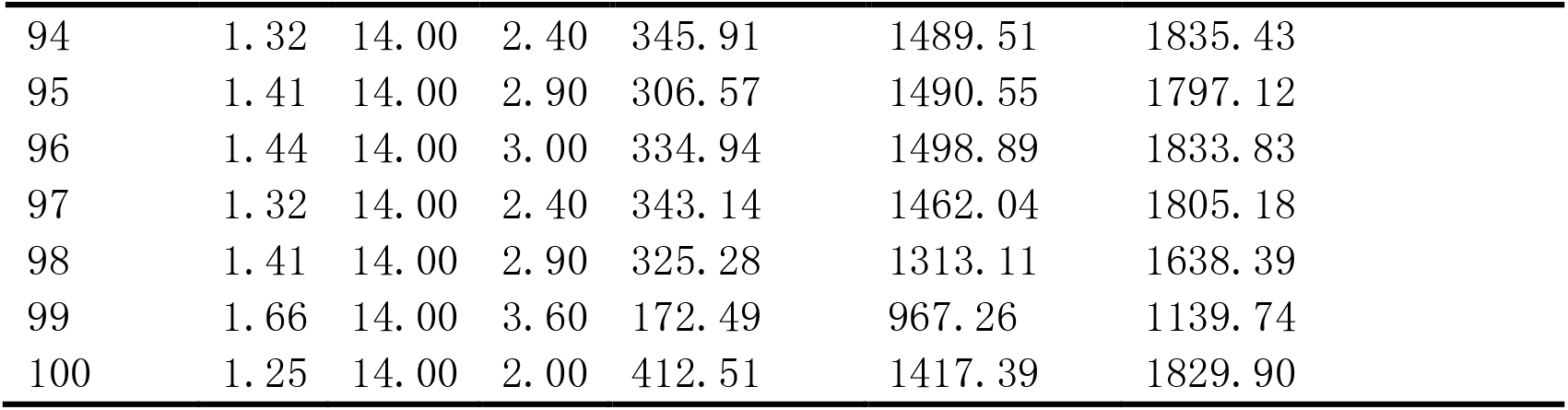
Distribution of the R_0_ (A), TI (B) and TE (C) for 100 random samplings

| Repeats | $R_0$ | TI | TE | RMSE<sub>(I)</sub> | RMSE<sub>(R)</sub> | RMSE<sub>(I)</sub> + RMSE<sub>(R)</sub> |
| --- | --- | --- | --- | --- | --- | --- |
| 1 | 1.54 | 14.00 | 3.30 | 232.06 | 1108.30 | 1340.36 |
| 2 | 1.29 | 14.00 | 2.30 | 328.27 | 1514.78 | 1843.05 |
| 3 | 1.29 | 14.00 | 2.30 | 278.94 | 1446.31 | 1725.26 |
| 4 | 1.41 | 14.00 | 2.90 | 325.89 | 1335.01 | 1660.90 |
| 5 | 1.51 | 14.00 | 3.20 | 259.15 | 1237.79 | 1496.94 |
| 6 | 1.57 | 14.00 | 3.40 | 235.95 | 1115.25 | 1351.20 |
| 7 | 1.44 | 14.00 | 3.00 | 336.74 | 1549.45 | 1886.20 |
| 8 | 1.54 | 14.00 | 3.30 | 276.54 | 1203.22 | 1479.76 |
| 9 | 1.36 | 14.00 | 2.50 | 230.20 | 1110.36 | 1340.56 |
| 10 | 1.44 | 14.00 | 3.00 | 328.24 | 1525.51 | 1853.75 |
| 11 | 1.44 | 14.00 | 3.00 | 333.88 | 1510.17 | 1844.05 |
| 12 | 1.26 | 14.00 | 2.20 | 239.48 | 1420.70 | 1660.18 |
| 13 | 1.47 | 14.00 | 3.10 | 344.07 | 1492.23 | 1836.30 |
| 14 | 1.41 | 14.00 | 2.90 | 324.17 | 1307.48 | 1631.65 |
| 15 | 1.41 | 14.00 | 2.90 | 265.93 | 1492.69 | 1758.62 |
| 16 | 1.29 | 14.00 | 2.30 | 329.13 | 1524.16 | 1853.29 |
| 17 | 1.46 | 14.00 | 3.10 | 307.72 | 1181.57 | 1489.29 |
| 18 | 1.41 | 14.00 | 2.90 | 273.56 | 1492.94 | 1766.51 |
| 19 | 1.51 | 14.00 | 3.20 | 257.96 | 1180.79 | 1438.75 |
| 20 | 1.26 | 14.00 | 2.20 | 258.77 | 1409.80 | 1668.56 |
| 21 | 1.46 | 14.00 | 3.10 | 312.57 | 1389.26 | 1701.83 |
| 22 | 1.46 | 14.00 | 3.10 | 319.22 | 1223.58 | 1542.81 |
| 23 | 1.43 | 14.00 | 3.00 | 294.43 | 1188.60 | 1483.03 |
| 24 | 1.44 | 14.00 | 3.00 | 330.24 | 1507.94 | 1838.18 |
| 25 | 1.69 | 14.00 | 3.70 | 199.45 | 955.89 | 1155.34 |
| 26 | 1.41 | 14.00 | 2.90 | 324.01 | 1293.32 | 1617.33 |
| 27 | 1.46 | 14.00 | 3.10 | 314.87 | 1391.49 | 1706.36 |
| 28 | 1.29 | 14.00 | 2.30 | 316.01 | 1523.79 | 1839.79 |
| 29 | 1.47 | 14.00 | 3.10 | 343.78 | 1523.28 | 1867.06 |
| 30 | 1.51 | 14.00 | 3.20 | 264.95 | 1224.86 | 1489.81 |
| 31 | 1.52 | 14.00 | 3.30 | 325.20 | 1370.43 | 1695.63 |
| 32 | 1.43 | 14.00 | 3.00 | 202.23 | 1271.15 | 1473.37 |
| 33 | 1.44 | 14.00 | 3.00 | 333.66 | 1541.24 | 1874.90 |
| 34 | 1.48 | 14.00 | 3.10 | 235.99 | 1250.52 | 1486.51 |
| 35 | 1.41 | 14.00 | 2.90 | 267.35 | 1492.76 | 1760.11 |
| 36 | 1.38 | 14.00 | 2.80 | 259.48 | 1443.04 | 1702.52 |
| 37 | 1.64 | 14.00 | 3.60 | 288.99 | 1200.59 | 1489.58 |
| 38 | 1.29 | 14.00 | 2.30 | 331.33 | 1278.37 | 1609.69 |
| 39 | 1.51 | 14.00 | 3.20 | 257.86 | 1201.83 | 1459.70 |
| 40 | 1.64 | 14.00 | 3.60 | 283.26 | 1191.24 | 1474.50 |
| 41 | 1.26 | 14.00 | 2.20 | 303.57 | 1533.43 | 1837.00 |
| 42 | 1.73 | 14.00 | 3.80 | 195.97 | 739.28 | 935.25 |
| 43 | 1.26 | 14.00 | 2.20 | 258.73 | 1421.11 | 1679.84 |
| 44 | 1.44 | 14.00 | 3.00 | 329.85 | 1548.18 | 1878.03 |
| 45 | 1.32 | 14.00 | 2.40 | 342.06 | 1462.74 | 1804.80 |
| 46 | 1.44 | 14.00 | 3.00 | 334.80 | 1508.12 | 1842.92 |
| 47 | 1.43 | 14.00 | 2.70 | 280.35 | 1183.74 | 1464.09 |
| 48 | 1.47 | 14.00 | 3.10 | 340.39 | 1494.90 | 1835.28 |
| 49 | 1.46 | 14.00 | 3.10 | 304.57 | 1312.61 | 1617.19 |
| 50 | 1.69 | 14.00 | 3.70 | 199.44 | 940.55 | 1139.99 |
| 51 | 1.64 | 14.00 | 3.60 | 280.98 | 1168.77 | 1449.75 |
| 52 | 1.52 | 14.00 | 3.30 | 341.47 | 1403.35 | 1744.83 |
| 53 | 1.41 | 14.00 | 2.90 | 254.30 | 1460.80 | 1715.11 |
| 54 | 1.25 | 14.00 | 2.00 | 352.72 | 1318.67 | 1671.39 |
| 55 | 1.51 | 14.00 | 3.20 | 238.67 | 1079.43 | 1318.10 |
| 56 | 1.44 | 14.00 | 3.00 | 329.99 | 1356.57 | 1686.57 |
| 57 | 1.29 | 14.00 | 2.30 | 333.46 | 1480.18 | 1813.64 |
| 58 | 1.47 | 14.00 | 3.10 | 336.68 | 1492.01 | 1828.69 |
| 59 | 1.41 | 14.00 | 2.90 | 309.87 | 1361.03 | 1670.90 |
| 60 | 1.38 | 14.00 | 2.80 | 298.04 | 1366.97 | 1665.01 |
| 61 | 1.41 | 14.00 | 2.90 | 267.13 | 1480.91 | 1748.03 |
| 62 | 1.40 | 14.00 | 2.90 | 291.41 | 1200.51 | 1491.92 |
| 63 | 1.41 | 14.00 | 2.90 | 260.20 | 1491.40 | 1751.60 |
| 64 | 1.26 | 14.00 | 2.20 | 312.96 | 1346.16 | 1659.12 |
| 65 | 1.29 | 14.00 | 2.30 | 330.69 | 1508.49 | 1839.17 |
| 66 | 1.47 | 14.00 | 3.10 | 342.46 | 1501.42 | 1843.88 |
| 67 | 1.43 | 14.00 | 3.00 | 301.38 | 1182.60 | 1483.98 |
| 68 | 1.44 | 14.00 | 3.00 | 327.74 | 1479.68 | 1807.42 |
| 69 | 1.41 | 14.00 | 2.90 | 325.82 | 1356.72 | 1682.54 |
| 70 | 1.41 | 14.00 | 2.90 | 303.12 | 1561.21 | 1864.33 |
| 71 | 1.44 | 14.00 | 3.00 | 336.34 | 1537.86 | 1874.20 |
| 72 | 1.44 | 14.00 | 3.00 | 325.27 | 1553.87 | 1879.14 |
| 73 | 1.47 | 14.00 | 3.10 | 346.19 | 1514.79 | 1860.98 |
| 74 | 1.49 | 14.00 | 3.20 | 324.28 | 1427.12 | 1751.40 |
| 75 | 1.47 | 14.00 | 3.10 | 339.75 | 1489.98 | 1829.73 |
| 76 | 1.44 | 14.00 | 3.00 | 329.81 | 1554.92 | 1884.73 |
| 77 | 1.46 | 14.00 | 3.10 | 312.38 | 1201.54 | 1513.92 |
| 78 | 1.49 | 14.00 | 3.20 | 327.08 | 1425.26 | 1752.34 |
| 79 | 1.41 | 14.00 | 2.90 | 308.17 | 1557.02 | 1865.19 |
| 80 | 1.44 | 14.00 | 3.00 | 333.98 | 1498.83 | 1832.81 |
| 81 | 1.41 | 14.00 | 2.90 | 319.86 | 1361.46 | 1681.32 |
| 82 | 1.46 | 14.00 | 3.10 | 313.13 | 1342.61 | 1655.74 |
| 83 | 1.26 | 14.00 | 2.20 | 298.31 | 1543.74 | 1842.04 |
| 84 | 1.52 | 14.00 | 3.30 | 343.84 | 1389.07 | 1732.91 |
| 85 | 1.51 | 14.00 | 3.20 | 204.54 | 1142.99 | 1347.54 |
| 86 | 1.29 | 14.00 | 2.30 | 334.83 | 1322.84 | 1657.67 |
| 87 | 1.47 | 14.00 | 3.10 | 342.76 | 1475.58 | 1818.34 |
| 88 | 1.41 | 14.00 | 2.90 | 307.41 | 1545.55 | 1852.96 |
| 89 | 1.46 | 14.00 | 3.10 | 315.48 | 1362.47 | 1677.95 |
| 90 | 1.41 | 14.00 | 2.90 | 313.66 | 1367.89 | 1681.55 |
| 91 | 1.32 | 14.00 | 2.40 | 349.91 | 1487.17 | 1837.08 |
| 92 | 1.32 | 14.00 | 2.40 | 345.09 | 1478.18 | 1823.28 |
| 93 | 1.29 | 14.00 | 2.30 | 329.34 | 1497.61 | 1826.94 |
| 94 | 1.32 | 14.00 | 2.40 | 345.91 | 1489.51 | 1835.43 |
| 95 | 1.41 | 14.00 | 2.90 | 306.57 | 1490.55 | 1797.12 |
| 96 | 1.44 | 14.00 | 3.00 | 334.94 | 1498.89 | 1833.83 |
| 97 | 1.32 | 14.00 | 2.40 | 343.14 | 1462.04 | 1805.18 |
| 98 | 1.41 | 14.00 | 2.90 | 325.28 | 1313.11 | 1638.39 |
| 99 | 1.66 | 14.00 | 3.60 | 172.49 | 967.26 | 1139.74 |
| 100 | 1.25 | 14.00 | 2.00 | 412.51 | 1417.39 | 1829.90 |

**Table 3.** Expected number of cases for infected, recovery and exposed

| Date | Infected | Recovery | Exposed | Date | Infected | Recovery | Exposed |
| --- | --- | --- | --- | --- | --- | --- | --- |
| 22-Jan | 425 | 28 | 426 | 7-Mar | 3121 | 185309 | 605 |
| 23-Jan | 537 | 58 | 532 | 8-Mar | 2542 | 186089 | 484 |
| 24-Jan | 676 | 97 | 664 | 9-Mar | 2068 | 186725 | 388 |
| 25-Jan | 849 | 145 | 829 | 10-Mar | 1680 | 187242 | 311 |
| 26-Jan | 1064 | 206 | 1034 | 11-Mar | 1364 | 187662 | 250 |
| 27-Jan | 1333 | 282 | 1290 | 12-Mar | 1106 | 188003 | 201 |
| 28-Jan | 1668 | 377 | 1607 | 13-Mar | 896 | 188279 | 161 |
| 29-Jan | 2084 | 496 | 1999 | 14-Mar | 726 | 188503 | 130 |
| 30-Jan | 2602 | 645 | 2482 | 15-Mar | 588 | 188685 | 104 |
| 31-Jan | 3243 | 831 | 3075 | 16-Mar | 476 | 188832 | 84 |
| 1-Feb | 4037 | 1062 | 3799 | 17-Mar | 385 | 188951 | 68 |
| 2-Feb | 5015 | 1351 | 4677 | 18-Mar | 311 | 189047 | 55 |
| 3-Feb | 6216 | 1709 | 5733 | 19-Mar | 252 | 189125 | 44 |
| 4-Feb | 7683 | 2153 | 6989 | 20-Mar | 203 | 189188 | 36 |
| 5-Feb | 9464 | 2702 | 8465 | 21-Mar | 164 | 189238 | 29 |
| 6-Feb | 11610 | 3377 | 10172 | 22-Mar | 133 | 189280 | 23 |
| 7-Feb | 14171 | 4207 | 12108 | 23-Mar | 107 | 189313 | 19 |
| 8-Feb | 14664 | 7750 | 17346 | 24-Mar | 87 | 189340 | 15 |
| 9-Feb | 16780 | 11416 | 22496 | 25-Mar | 70 | 189361 | 12 |
| 10-Feb | 20084 | 15611 | 27606 | 26-Mar | 57 | 189379 | 10 |
| 11-Feb | 24265 | 20632 | 32449 | 27-Mar | 46 | 189393 | 8 |
| 12-Feb | 29015 | 26698 | 36597 | 28-Mar | 37 | 189404 | 6 |
| 13-Feb | 33960 | 33952 | 39545 | 29-Mar | 30 | 189414 | 5 |
| 14-Feb | 38652 | 42442 | 40874 | 30-Mar | 24 | 189421 | 4 |
| 15-Feb | 42614 | 52105 | 40409 | 31-Mar | 19 | 189427 | 3 |
| 16-Feb | 45430 | 62758 | 38286 | 1-Apr | 16 | 189432 | 3 |
| 17-Feb | 46835 | 74116 | 34898 | 2-Apr | 13 | 189436 | 2 |
| 18-Feb | 46759 | 85824 | 30762 | 3-Apr | 10 | 189439 | 2 |
| 19-Feb | 45323 | 97514 | 26371 | 4-Apr | 8 | 189442 | 1 |
| 20-Feb | 42782 | 108845 | 22107 | 5-Apr | 7 | 189444 | 1 |
| 21-Feb | 39456 | 119540 | 18213 | 6-Apr | 5 | 189445 | 1 |
| 22-Feb | 35663 | 129404 | 14809 | 7-Apr | 4 | 189447 | 1 |
| 23-Feb | 31684 | 138320 | 11925 | 8-Apr | 4 | 189448 | 1 |
| 24-Feb | 27738 | 146241 | 9537 | 9-Apr | 3 | 189449 | 0 |
| 25-Feb | 23982 | 153175 | 7591 | 10-Apr | 2 | 189449 | 0 |
| 26-Feb | 20517 | 159171 | 6025 | 11-Apr | 2 | 189450 | 0 |
| 27-Feb | 17396 | 164300 | 4774 | 12-Apr | 1 | 189451 | 0 |
| 28-Feb | 14639 | 168649 | 3781 | 13-Apr | 1 | 189451 | 0 |
| 29-Feb | 12239 | 172309 | 2995 | 14-Apr | 1 | 189451 | 0 |
| 1-Mar | 10178 | 175369 | 2374 | 15-Apr | 1 | 189451 | 0 |
| 2-Mar | 8424 | 177913 | 1883 | 16-Apr | 1 | 189452 | 0 |
| 3-Mar | 6946 | 180019 | 1496 | 17-Apr | 1 | 189452 | 0 |
| 4-Mar | 5708 | 181756 | 1191 | 18-Apr | 0 | 189452 | 0 |
| 5-Mar | 4678 | 183183 | 949 | 19-Apr | 0 | 189452 | 0 |
| 6-Mar | 3825 | 184352 | 757 | 20-Apr | 0 | 189452 | 0 |

**Fig 3.**
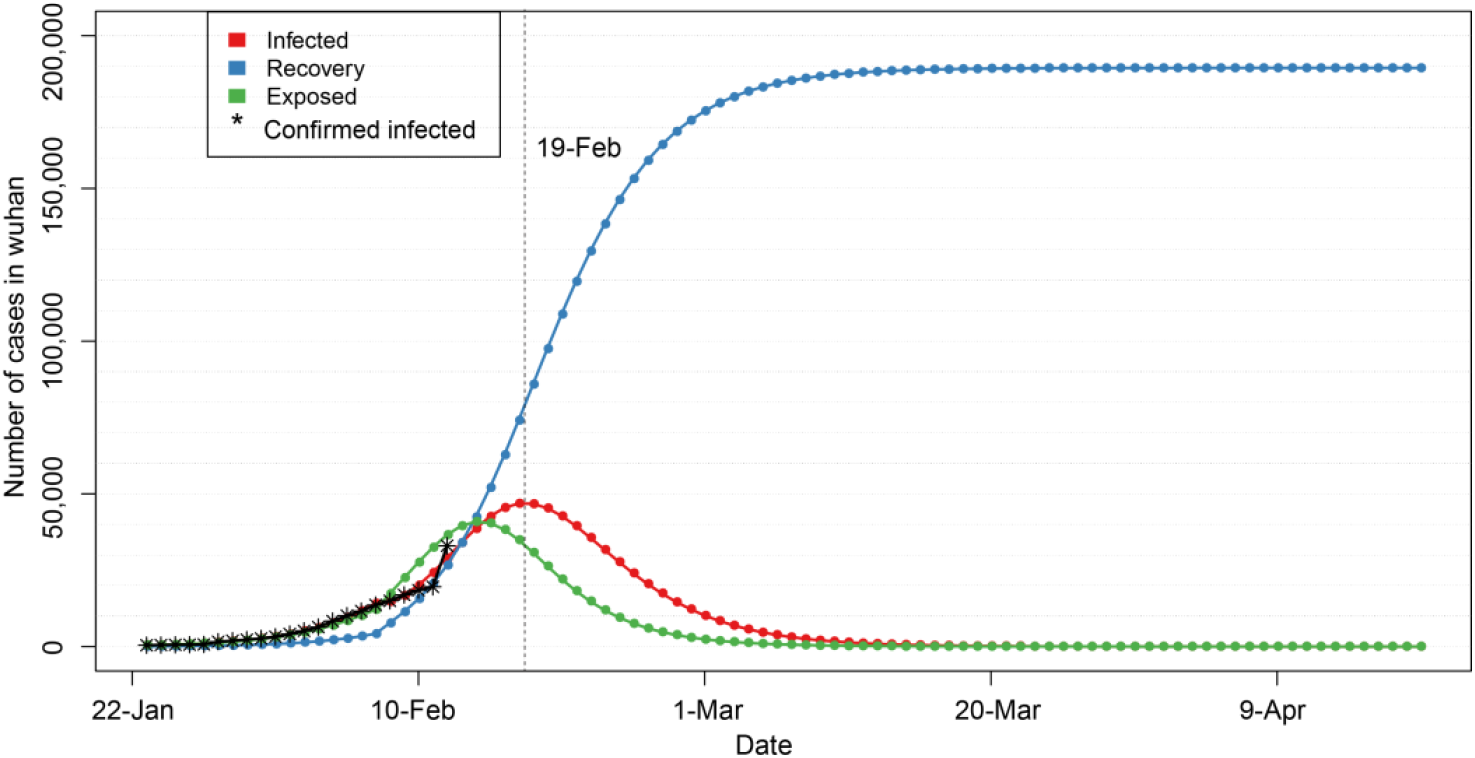
Epidemic trend of 2019-nCoV within 90 days after the closure of Wuhan.

In Figure 3 and Figure 4, red line indicates the trend of cumulative infection number over time, blue line is the trend of cumulative rehabilitation number over time, and green line is the trend of cumulative latent number over time. Vertical dash line indicates the peak time of cumulative infection number.

**Fig 4.**
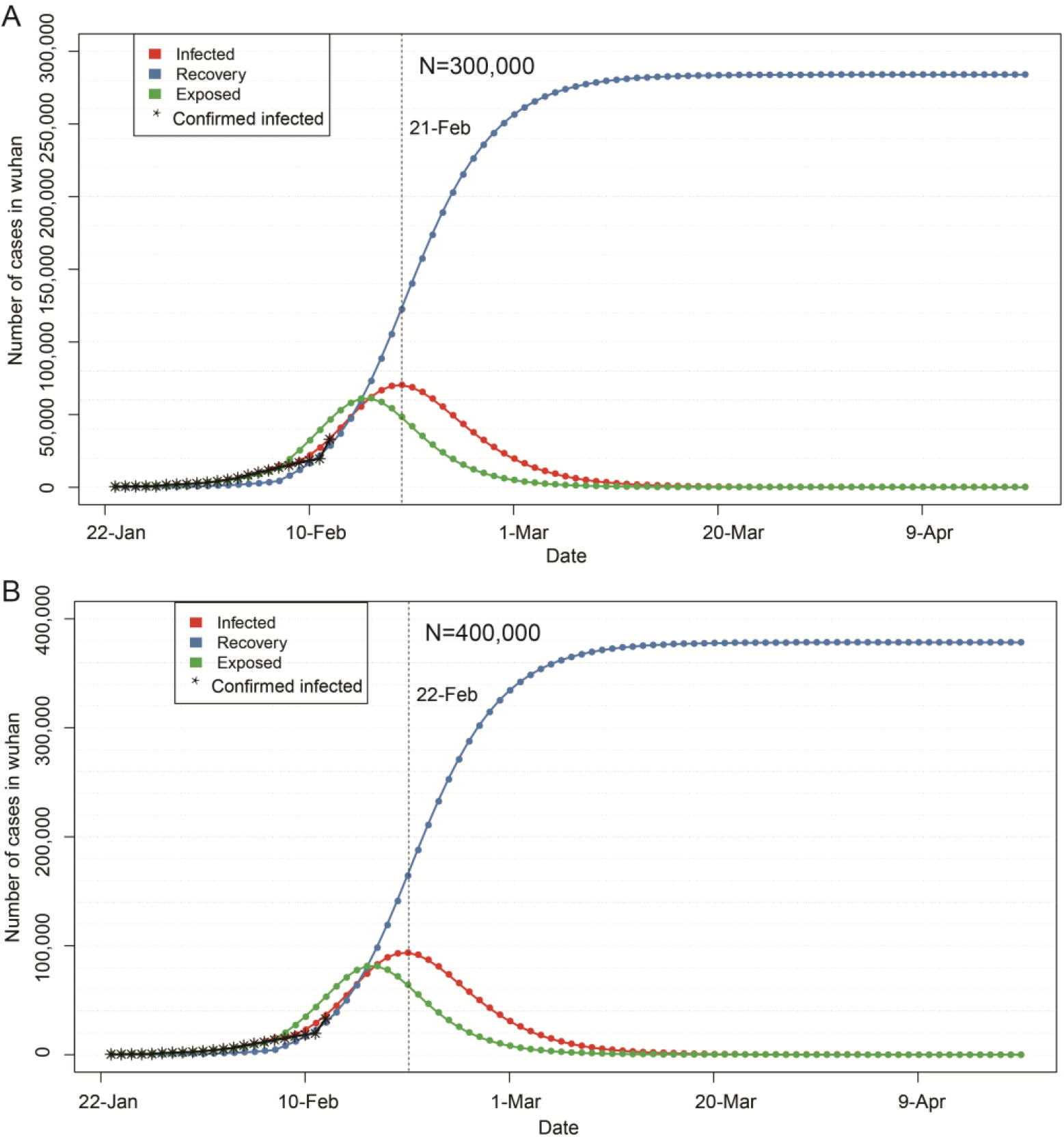
Epidemic trend of 2019-nCoV within 90 days after the closure of Wuhan city assuming the number of susceptible people n = 300000 (A) and 400000 (B)

If the epidemic situation is not properly controlled, the number of susceptible population will continue to increase on the basis of current N = 200000. If the number of susceptible population increases to N = 300000, and other parameters remain unchanged, the peak number can increase to 75000, and the epidemic peak time will also be postponed at around February 21 (Figure 4). If it is increased to N = 400000 and other parameters remain unchanged, the peak number can be increased to 100000, and the epidemic peak will be postponed to around February 22 (Figure 4). Even in both cases, the epidemic would subside in early March, and disappear gradually towards the late March.

## Discussion

Although some modeling studies on the epidemiological characteristics of 2019-nCoV epidemic have been reported so far, they had some limitations, such as the data come from the early stage of the epidemic. Due to the rapid change of the epidemic situation and the closure of Wuhan on January 23, many parameters related to the model have also changed, which affect the applicability and reliability of the model. This study used the latest 2019 nCoV data in Wuhan area, analyzed the epidemiological characteristics of 2019 nCoV epidemic after in Wuhan city was shut down. Compared with other studies, the R_0_ value produced in this study is smaller, indicating that the closure and subsequent measures have played an important role in the spread of the epidemic.

The infection time index (TI) obtained in this study was higher than that of SARS[12] and MERS[13], but lower than that of 2019-nCoV in literatures[14] reported earlier. This result may be related to the sudden outbreak of the epidemic, the lack of medical resources for early response, and the failure of timely diagnosis and treatment of infected patients. A large number of mild patients and asymptomatic virus carriers were not isolated in time. The incubation period (TE) is about 3 days, which is close to the data in the reference^[14]^.

According to the latest reported data, the cumulative number of people infected on February 13 and 14 was 35991 and 37914 respectively, which is close to the number predicted by our estimation (Table 2). According to this study, the number of infected people will reach the peak in February 19 at about 45000 infected individuals.

## Conclusions

With the implementation of more follow-up measures, including strict restrictions on people going out, accelerating the treatment of infected individuals, and clinical trials of new drugs, the development of 2019-nCoV epidemic in Wuhan will be effectively controlled, and the number of infected individuals will gradually decrease. It was expected that the epidemic would subside in early March, and disappear gradually towards the late March. If the epidemic situation is not properly controlled, the peak of infected number can be further increased and the peak time will be a little postponed.

## Data Availability

The data were collected from the official website of Hubei Provincial Health Committee (http://wjw.hubei.gov.cn/)

## Declarations

### Ethics approval and consent to participate

The ethical approval or individual consent was not applicable.

### Availability of data and materials

All data and materials used in this work were publicly available.

### Consent for publication

Not applicable.

### Disclaimer

The funding agencies had no role in the design and conduct of the study; collection, management, analysis, and interpretation of the data; preparation, review, or approval of the manuscript; or decision to submit the manuscript for publication.

